# Modeling the Spring 2020 New York City COVID-19 Epidemic: New Criteria and Methods for Prediction

**DOI:** 10.1101/2020.06.12.20130005

**Authors:** D. A. Barlow, J. K. Baird

## Abstract

We report here on results obtained using the SIR epidemic model to study the spring 2020 COVID-19 epidemic in New York City (NYC). An approximate solution is derived for this non-linear system which is then used to derive an expression for the time to maximum infection. Additionally, expressions are obtained for estimating the transmission and recovery parameters using data collected in the first ten days of the epidemic. Values for these parameters are then generated using data reported for the spring 2020 NYC COVID-19 epidemic which are then used to estimate the time to maximum infection and the maximum number of infected. Complete details are given so that the method can be used in the event of future epidemics. An additional result of this study is that we are able to suggest a unique mitigation strategy.

## 1 Introduction

Mathematical analysis of the spread of pathogens in the environment, whether amongst plants, animals or humans, has been an active area of research since the seminal paper of Kermack and McKendrik in 1927 [1, 2]. In this work they provide a general description for an epidemic via a system of differential equations. They go on to use these to study an early twentieth century plague epidemic. More recently, similar kinetic models have been used to study other infectious outbreaks in human and animal populations including cholera [3, 4], anthrax [5, 6], SARS [7], H1N1 [8] and chlamydia [9] to name a few.

The Kermack-McKendrik, or so called, SIR model, relates the number of susceptible individuals *S*, number of infected persons *I*, with the number of recovered *R* through a system of three differential equations:

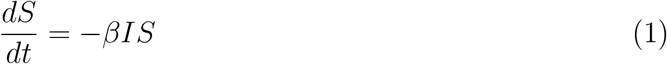

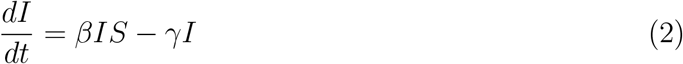

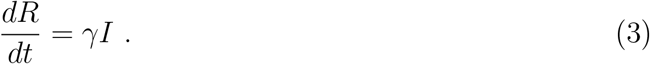

Here *t* represents time, *β* is the specific transmission coefficient and *γ* the recovery coefficient. In this work both *β* and *γ* are assumed to remain constant. Eq. (2) results from the fact that

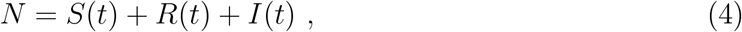

where *N* is the total number of persons in the single-compartment, homogeneous set.

Even though, with *β* and *γ* held constant, the SIR model is one of the more simple epidemic models this non-linear system has defied closed-form analytical solution and is thus routinely solved numerically. Recently, an exact solution in parametric form was reported by Harko et al. [10]. Given initial values for *S, R* and *I*, and values for the parameters *β* and *γ*, the result of Harko et al. is shown to equivalent to the numerical one. However, values for *β* and *γ* before the onset or during the early stages of an epidemic are not always available especially in the case of a novel pathogen as with COVID-19 in the spring of 2020.

In this report we present results from using the Kermack-McKendrick model to study the 2020 spring COVID-19 outbreak in NYC. A scheme is given for estimating *β* and *γ* using testing data from the early stage of an epidemic. An approximate solution for the entire system of Eqs. (1) through (3) leads to an estimate for the time to maximum infection, *t*_*p*_, in terms of *β, γ* and *S*_*o*_ where *S*_*o*_ is the number of susceptible people at *t* = 0. It is then shown how these parameters can be used within expressions previously reported [7, 11, 12] for the maximum number of infected persons, *I*_*p*_, and the final number of susceptible though not infected individuals *S*_*∞*_, to compute estimates for these quantities. Finally, the estimated values for *β* and *γ* are used to assemble the curves for *S*(*t*), *I*(*t*) and *R*(*t*), both numerically and using the approximate solution, for the case of the spring 2020 COVID-19 epidemic in NYC. The approach is general and may be used to estimate *β* and *γ* for other similar epidemics in the future.

The effects of mitigation are studied by viewing a three-dimensional plot of *I* vs. *β* and time. This plot reveals how *flattening the curve* in two-dimensions is equivalent to *bending the ridge* in three. The result being that as *β* is decreased through mitigation the peak of the ridge, *I*_*p*_, is lowered but the epidemic takes a path through finite *I*(*t*) that becomes longer in time eventually becoming infinite. It is further suggested here that by using a particular mitigation approach, and thus taking a particular route through *I*(*β, t*) space, that the intensity and time span of the epidemic may be lessened.

## 2 Estimating *β* and *γ* using early time data

In this section we derive approximate expressions for the parameters *β* and *γ* that are given solely in terms of time dependent data for *I* taken early in the course of the epidemic. We have the initial conditions *S*(0) = *S*_*o*_, *R*(0) = 0 and *I*(0) = *I*_*o*_.

We consider only the situation where *βS*_*o*_ *> γ*. At the earliest times in the epidemic we claim that Eq. (2) will be dominated by the first term on the right so that we can write the following approximation for Δ*I/*Δ*t*:

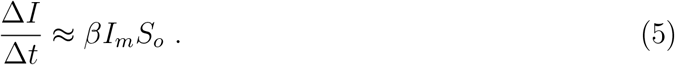

Here *I*_*m*_ is the mean value for *I* over the time interval Δ*t*. Solving this for *β* we get the formula used to estimate this parameter

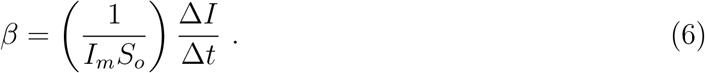

Now, consider some time interval Δ*t*_1_ later in the *I* vs. *t* data but still relatively early in the epidemic, say within the first 10 days. Then the slope of Δ*I/*Δ*t*_1_ over this interval can be approximated using Eq. (5) using the value for *β* estimated with the earlier time interval Δ*t*. However, this would over estimate Δ*I/*Δ*t*_1_ as the second term in Eq. (2) can not be ignored at this later time. We conjecture then that a better estimate for the slope over Δ*I/*Δ*t*_1_ be given by

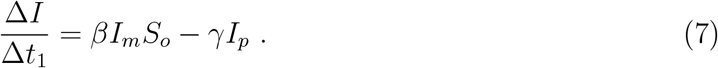

where *I*_*p*_ is the peak value for *I*. Solving for *γ* yields

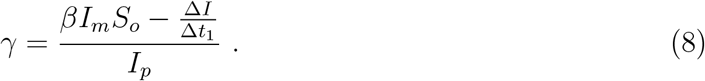

Since *I*_*p*_ will be unknown we use the approximation that *I*_*p*_ ≈ *S*_*o*_. It will be demonstrated in a later section how Eqs. (6) and (8) can be used along with early time data for *I* vs. time from a real epidemic to estimate *β* and *γ*.

## 3 Approximate Solution for the Kermack-McKendrick Equations

In this work we consider the epidemic, modeled by Eqs. (1) through (3), where *βS*_*o*_*/γ >* 1. From a theorem due to Hethcote [7, 11] this implies several things. First, that *I*(*t*) will have a positive maximum which is labeled *I*_*p*_ and then *I →* 0 as *t → ∞*. Secondly, *I*_*p*_ can be given by

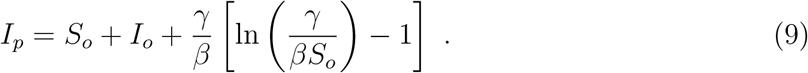

Additionally, the remaining number of susceptible people not infected at the end of the epidemic, *S*_*∞*_, is given by

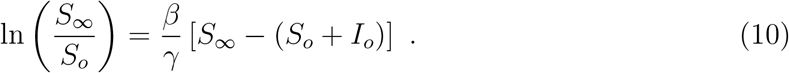

A useful approximate solution for Eqs. (1) through (3) can be arrived at starting with the integrated version of Eq. (3)

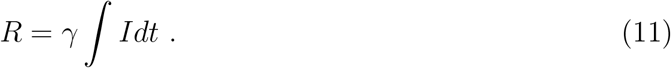

Eqs. (1) and (3) can be used to derive

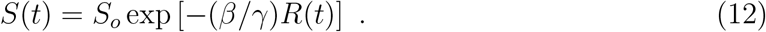

Using Eq. (11) in Eq. (12) we have

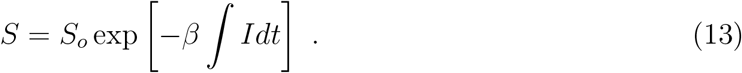

Using Eqs. (11) and (13) in Eq. (4) gives

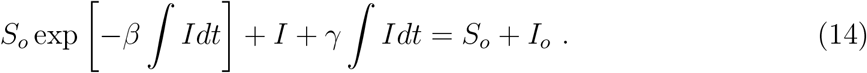

Operating on both sides of eq. (14) with *d/dt* yields

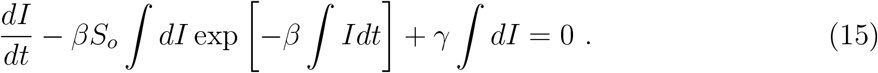

We let ∫ *dI* = *I* + *c*_1_, where *c*_1_ is some constant, and the above becomes

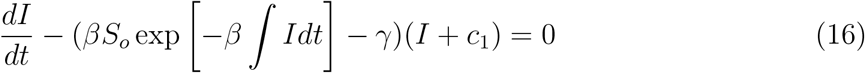

*∫ Idt* is some function of time, that is, *R*(*t*). We speculate that during the period of time over which *I* is significant *R* can be modeled as being quadratic with respect to time so we let ∫ *Idt* = *kt*^2^ where *k* is some yet to be determined parameter. With these replacements in Eq. (16) we have the separable situation:

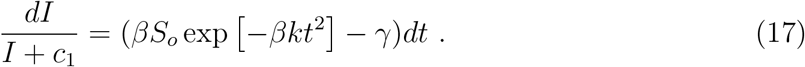

Integrating both sides of Eq. (17) and solving for *I* we get

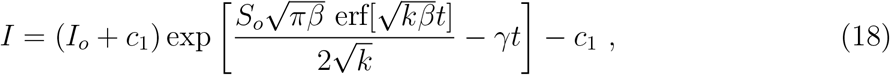

where erf denotes the error function. There is the final condition that *I →* 0 as *t → ∞* so it must be that *c*_1_ = 0. The final expression for *I* is then given by

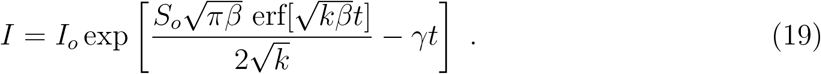

More can be learned about *k* by considering the time of maximum *I* which we label as *t*_*p*_. We conclude that *I* will be a maximum when the argument of the exponential function in Eq. (19) is a maximum. Computing the first time derivative of this yields

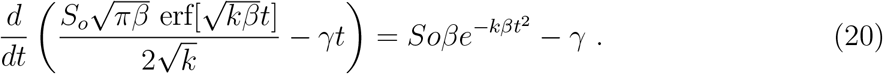

Setting this equal to zero we solve for the time to maximum infection, *t*_*p*_:

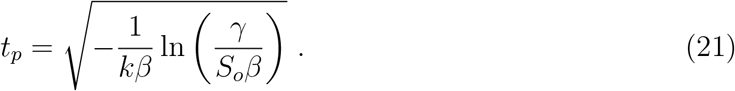

Therefore, it must be that *γ < S*_*o*_*β*. Using Eq. (21) in eq. (19) we are lead to an approximate expression for *I*_*p*_:

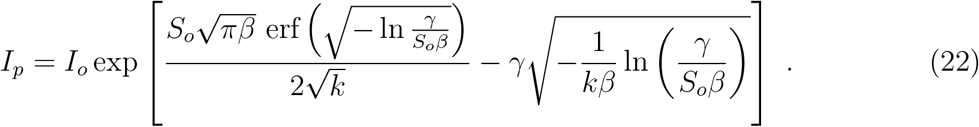

This is equated to the true expression for *I*_*p*_ given by Eq. (9):

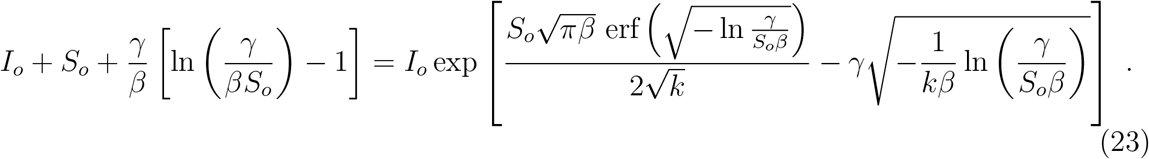

Solving the above for *k* we get

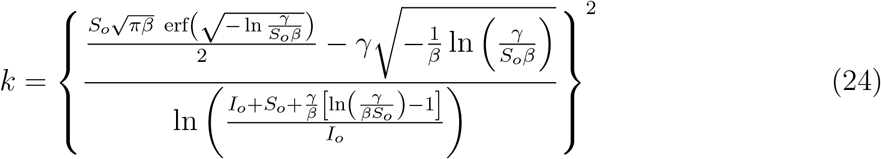

With values for *β, γ, S*_*o*_ and *I*_*o*_, *I*_*p*_ and *k* can be computed and thus *t*_*p*_ estimated. Also, the approximate curve for *I*(*t*) can be generated and plotted.

Now, *S*(*t*) and *R*(*t*) can be estimated by using Eq. (19) in Eqs. (3) and (12). The integral of Eq. (19) must be computed numerically. For purposes of simplification we propose the use of the following tabulated function

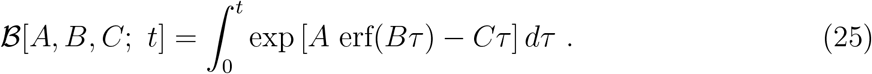

Using this notation, along with Eq. (3), with we have that

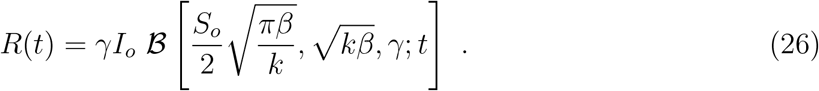

Eq. (26) in Eq. (12) gives the approximation for *S*(*t*),

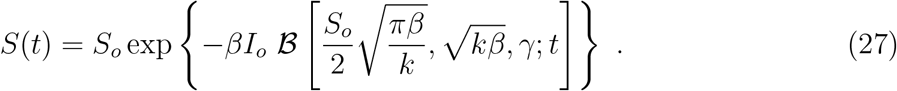

## 4 Analysis of Epidemic Data

Recently the NYC Department of Health released data on the NYC spring 2020 COVID-19 epidemic [13]. These data included a daily tally of the percentage of positive test results and a daily three-day average of percentage positive tests results. We take the positive test result as a measure of the number of the infected people per day that is, *I* vs. time. Additionally, data was provided for the daily number of hospitalizations and cases during the same time period. This data was given for an approximately 80 day period starting in early March 2020. The first positive test percentage reported was for 3 March while the first three-day average was on 5 March. In the report on number of hospitalizations it is listed that the first case was reported on 29 February. Therefore, 29 February will be taken as the zero point for time in the epidemic. Here we will analyze the data on positive test results given as a three day average, that is, reported daily for the previous three days. It is demonstrated in this section how data from the first ten days of this data can be used to estimate values for the constants *β* and *γ*.

We consider an example set of 1000 susceptible people. The epidemic is assumed to be started by one individual so that *I*_*o*_ = 1 and *S*_*o*_ = 999. That is, we assume that 0.1% of 1000 people were infected when data collection commenced. We assume that the percentage of positive test results applies to this entire set. The first three day average of 0.09 percent was reported on 5 March 2020. With the epidemic starting on 29 February we set Δ*t* = 4.0 days. Therefore the infected persons after the first four day period must be *I* = (0.09)(1000) = 90 so that Δ*I* = 89. *I*_*m*_ is the mean value over this time period. This data is now used in Eq. (6) to estimate *β* and the result is listed in Table 1.

**Table 1:** Parameters computed in this SIR study of the spring 2020 NYC COVID-19 epidemic. *-taken from Ref. [13]. ^*♣*^ -from the curve fit of *I* shown in Fig. 4 taken from the numerical solution to Eqs. (1), (2) and (3). By Eq. (10) *S*_*∞*_ *≈* 0 in this case.

| $I_p$ | $t_p$ (days) | $\beta$ (days)$^{-1}$ | $\gamma$ (days)$^{-1}$ | $R_o$ | $k$ (days)$^{-2}$ |
| --- | --- | --- | --- | --- | --- |
| 695.1 | 28.5 | $4.95 \times 10^{-4}$ | 0.04419 | 11.2 | 6.0 |
| 700* | 27.0* | $3.85 \times 10^{-4}$ ♣ | 0.0365 ♣ | 10.53 ♣ | |

Now a later time period, Δ*t*_1_ is considered and Eq. (8) used to estimate *γ*. For three time intervals, beyond day four, *γ* will be computed using Eq. (8) for each and then a mean value determined. For day six to seven the percentage increases from 0.06 to 0.1. For day seven to eight the change is 0.1 to 0.12. Finally, we consider data from day eight to nine where the percentage increased from 0.12 to 0.19. In each case Δ*t*_1_ = 1.0 day. Since *I*_*p*_ is not yet known we assume that the epidemic is significant and assume that *I*_*p*_ *≈ S*_*o*_. The mean of these three values is listed in Table 1. Additionally, a quantity often computed during studies of this type, the so-called reproduction number *R*_*o*_ where *R*_*o*_ = *βS*_*o*_*/γ*, is computed and listed along with the above mentioned quantities in the first row of Table 1.

Finally, our estimated values for *β* and *γ* can be used in Eq. (24) to estimate the parameter *k* then Eqs. (19), (26) and (27) can be used to yield the full curves for *I*(*t*), *R*(*t*) and *S*(*t*) for the COVID-19 spring 2020 NYC epidemic. These curves are depicted in Figures 1, 2 and 3. In Fig. 4 the numerical solution for Eqs. (1), (2) and (3) was fitted to the data shown in Fig. 1 by adjusting the values for *β* and *γ*. Values from this fit are listed in the second row of Table 1.

**Figure 1:**
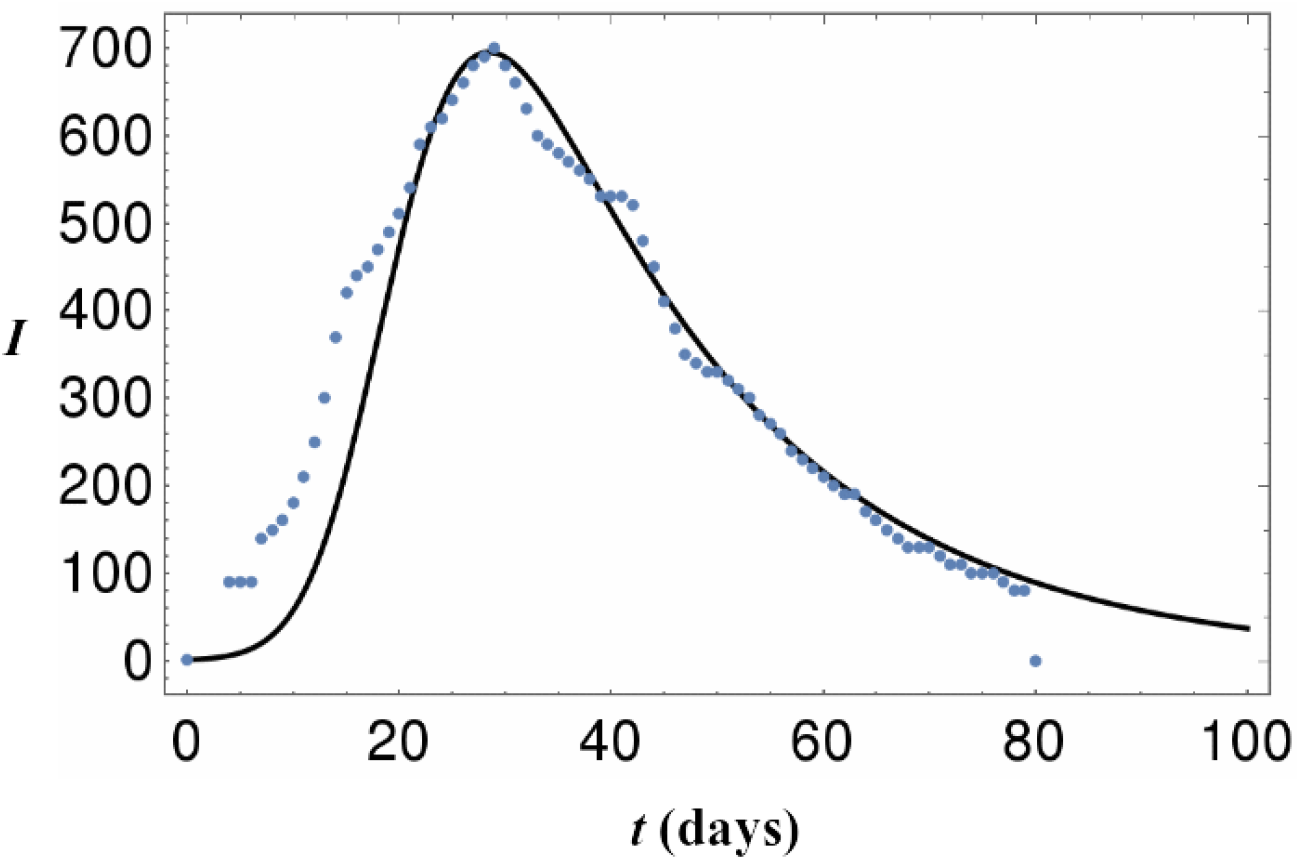
*I* vs. time. Data points are percentage of 1000 persons testing positive based upon data from Ref. [13]. Solid curve as estimated by Eq. (19). *β* and *γ* from the first row of Table 1, generated using the prediction scheme outlined in this report, were used.

**Figure 2:**
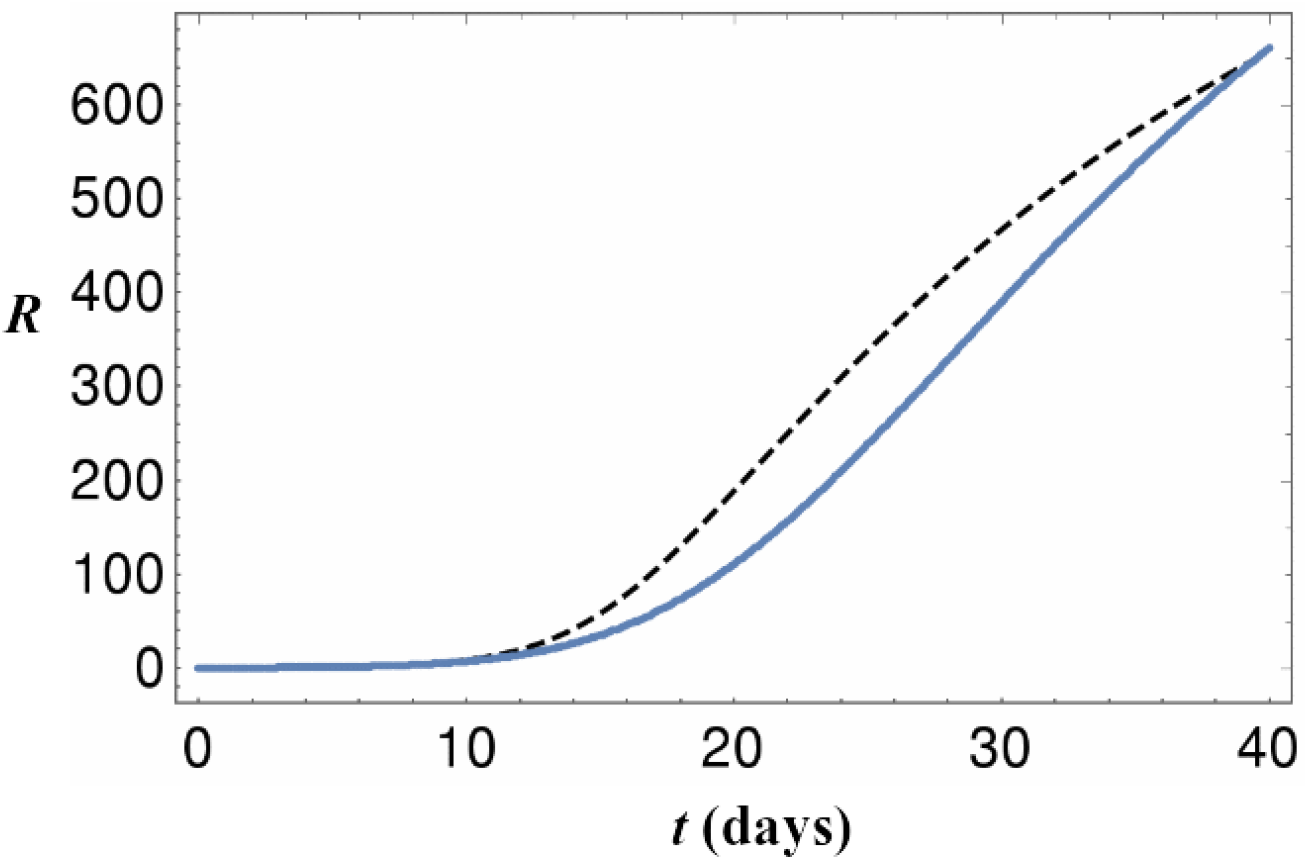
*R* vs. time. Solid curve as estimated by Eq. (26). Dashed curve is the numerical solution from of Eqs. (1), (2) and (3). In both cases *β* and *γ* from the first row of Table 1 were used.

**Figure 3:**
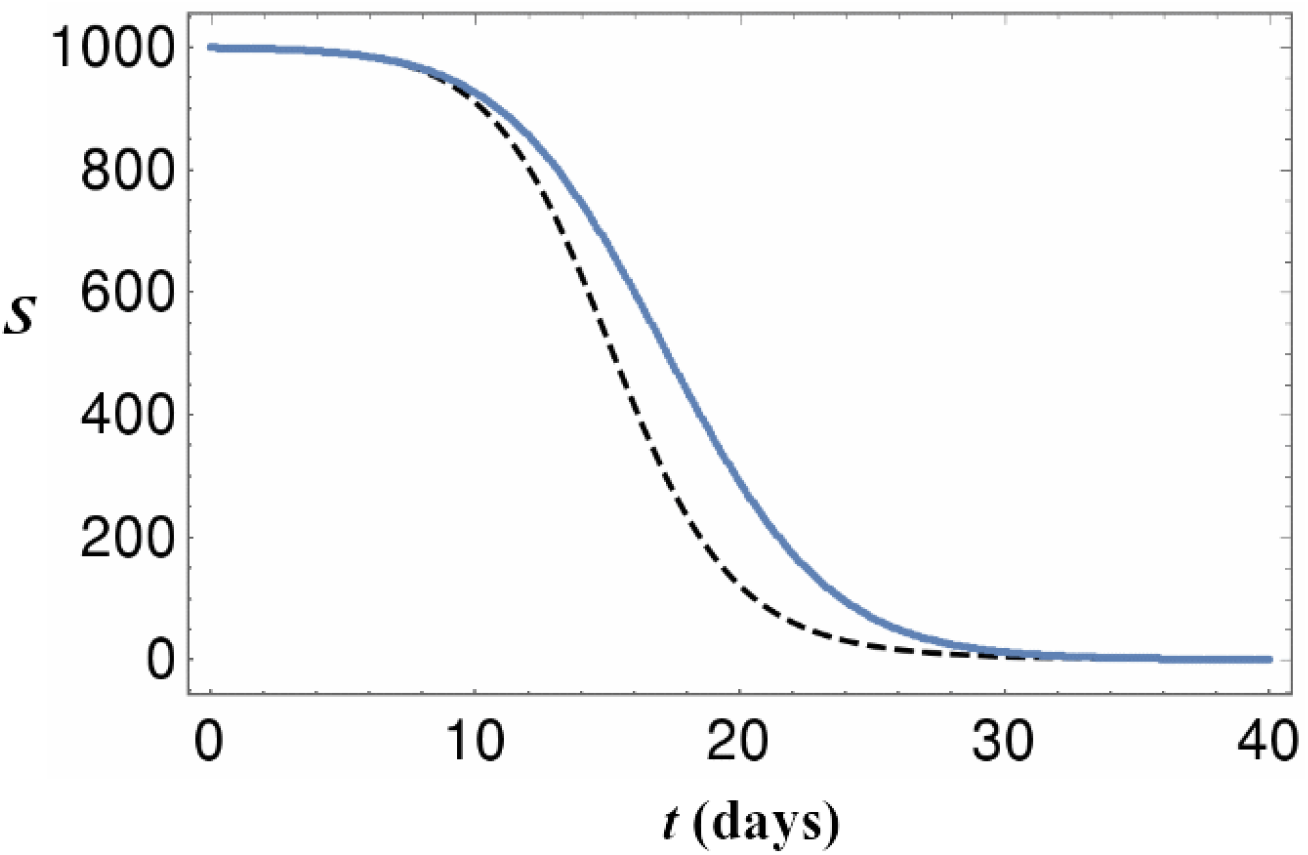
*S* vs. time. Solid curve as estimated by Eq. (27). Dashed curve is the numerical solution from of Eqs. (1), (2) and (3). In both cases *β* and *γ* from the first row of Table 1 were used.

**Figure 4:**
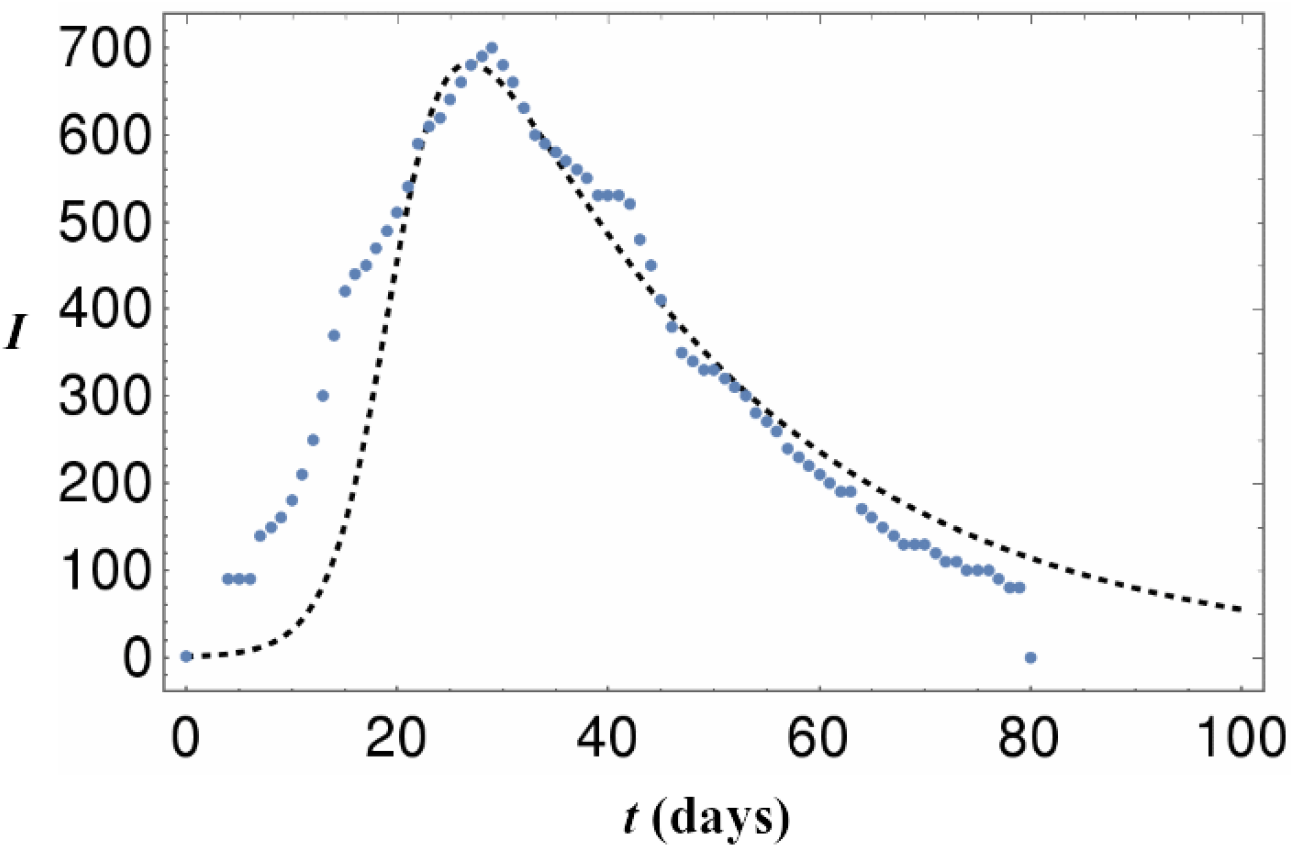
*I* vs. time. Dashed curve is the numerical solution from of Eqs. (1), (2) and (3). *β* and *γ* from the second row of Table 1 were used.

## 5 Discussion

In this report it was shown how early-time epidemic data can be used to predict two important parameters used in the SIR model. Using daily percentage testing positive data reported for the spring 2020 COVID-19 epidemic in NYC as an example, the scheme is employed to predict the number of infected persons at the peak of infection and the time from the start of the epidemic until peak infection. Further, the general SIR curves for *I*(*t*), *R*(*t*) and *S*(*t*) are approximated and shown to be a reasonable match to data or the curves from the numerical result.

From the data in Table 1 it is seen that the model using the values for *β* and *γ* generated using early-time data, predicts a value for the number of maximum infected people to within about 1.0 % of the reported value. The predicted time to peak infection is within about 6% of the reported time. These errors could be considered significant for a theoretical model being used to describe some physical phenomena or system. However, for the purpose of predicting the time dependent nature of numbers of infected, recovered and susceptible people during an epidemic, these errors should not preclude such a model from being significantly useful to the forecaster.

The value for *β* and *R*_*o*_ determined in this study are indicative of a pathogen that spreads relatively quickly and efficiently. Some caution is warranted when quoting this value however. Though it is expected that the COVID-19 virus is in fact more easily transferable than the common flu, we have assumed here the group that was tested was typical of the entire population, an assumption that may or may not be accurate. Additionally, the reproduction number in this model is sensitive to the choice of value for the ratio *I*_*o*_*/*(*S*_*o*_ + *I*_*o*_). In this work we selected 1/1000 for this ratio, that is to say that the epidemic started by having one infected person per 1000 susceptible people, or at least, that was the situation when data collection commenced. Interestingly, as this ratio decreases, the reproduction number increases thus indicating that there were likely more than 0.1% infected at the time the NYC data collection commenced as COVID-19 reproduction numbers are speculated to be in the neighborhood of 4.0 [2].

Of course if *R*_*o*_ *<* 1 the infection never spreads and the epidemic dies out. Kermack and McKendrick consider this in terms of a so called population density given by *γ/β*, [1]. If the population density is less than *γ/β* the epidemic dies out whereas it spreads and reaches a maximum *I* for cases where the population density exceeds *γ/β*. Using *β* and *γ* from the first row of Table 1 we get *γ/β* ≈ 89 thus indicating that for these values of *β* and *γ* at least 89 persons are required for the epidemic to develop. This would in fact be the minimum required population for any case where the ratio *I*_*o*_*/*(*S*_*o*_ + *I*_*o*_) = 1*/*1000 as *I*_*o*_ ≥1.

Much has been said recently about the effect of mitigation on the shape of the curve for *I*(*t*). It has been assumed that by the adoption of aggressive mitigation techniques and practices the transfer rate *β* can be lowered. This has the effect of lowering the peak value for *I* and spreading the epidemic out over a longer time period. This change is referred to as *flattening the curve* [14]. It is interesting to observe the effect on *I* of varying *β* through a 3D plot of *I* vs. *t* and *β* at constant *γ*. Using the predicted value for *γ* from the first row of Table 1, along with Eqs. (24) and (19), a plot for *I*(*β, t*) is generated over a range of values for *β* that includes those found in this study for the NYC epidemic. This plot is shown in Fig. 4.

It is seen from this figure that as *β* is decreased the peak for *I* lessens and shifts towards later times. This change is seen as a flattening of the curve in 2D but becomes a *bending of the ridge* in 3D. This effect is seen more clearly via a top view as depicted in Fig. 5.

**Figure 5:**
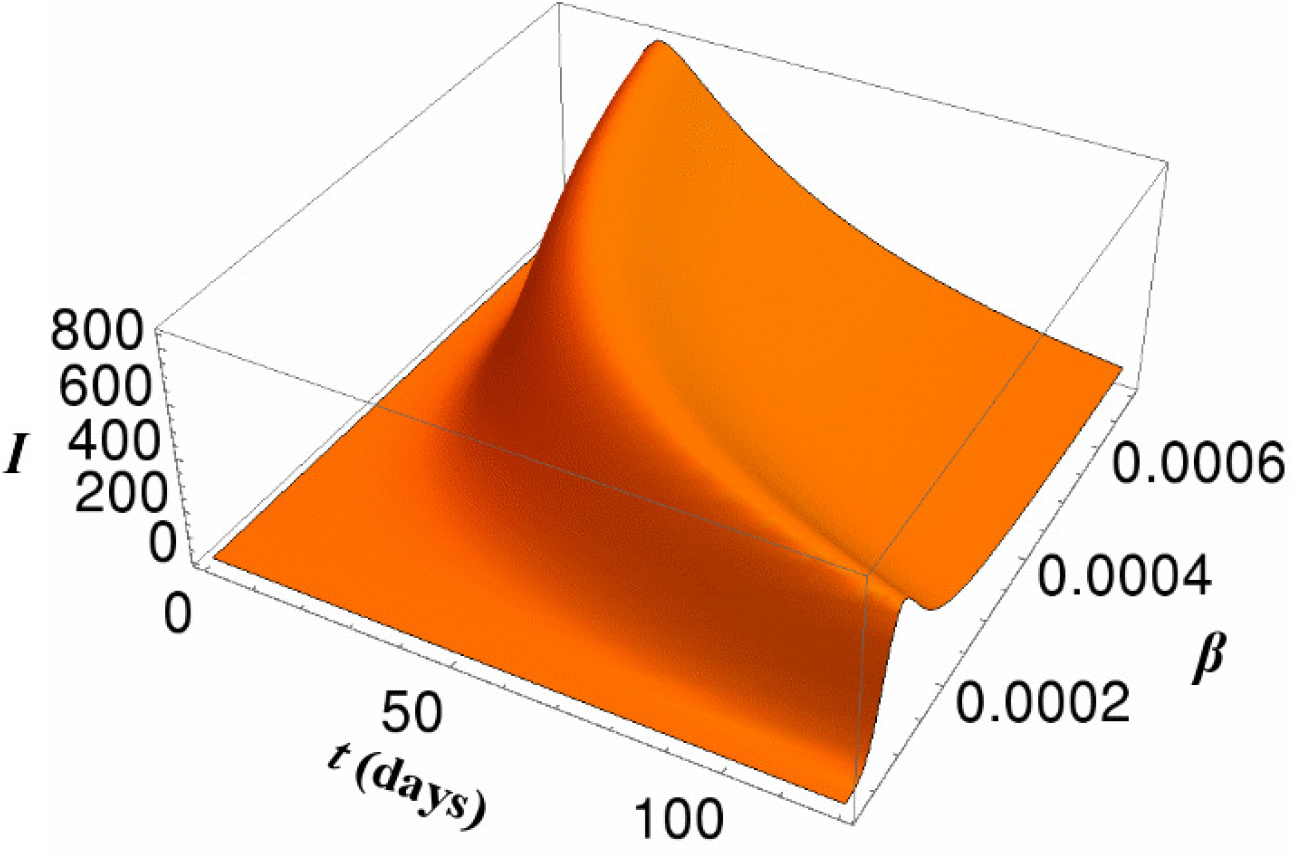
*I* vs. *β* and time at constant *γ*. Here *γ* from the first row of Table 1 was used. The surface is given by using Eq. (24) in (19). *β* has units of days^−1^ per 1000 people.

**Figure 6:**
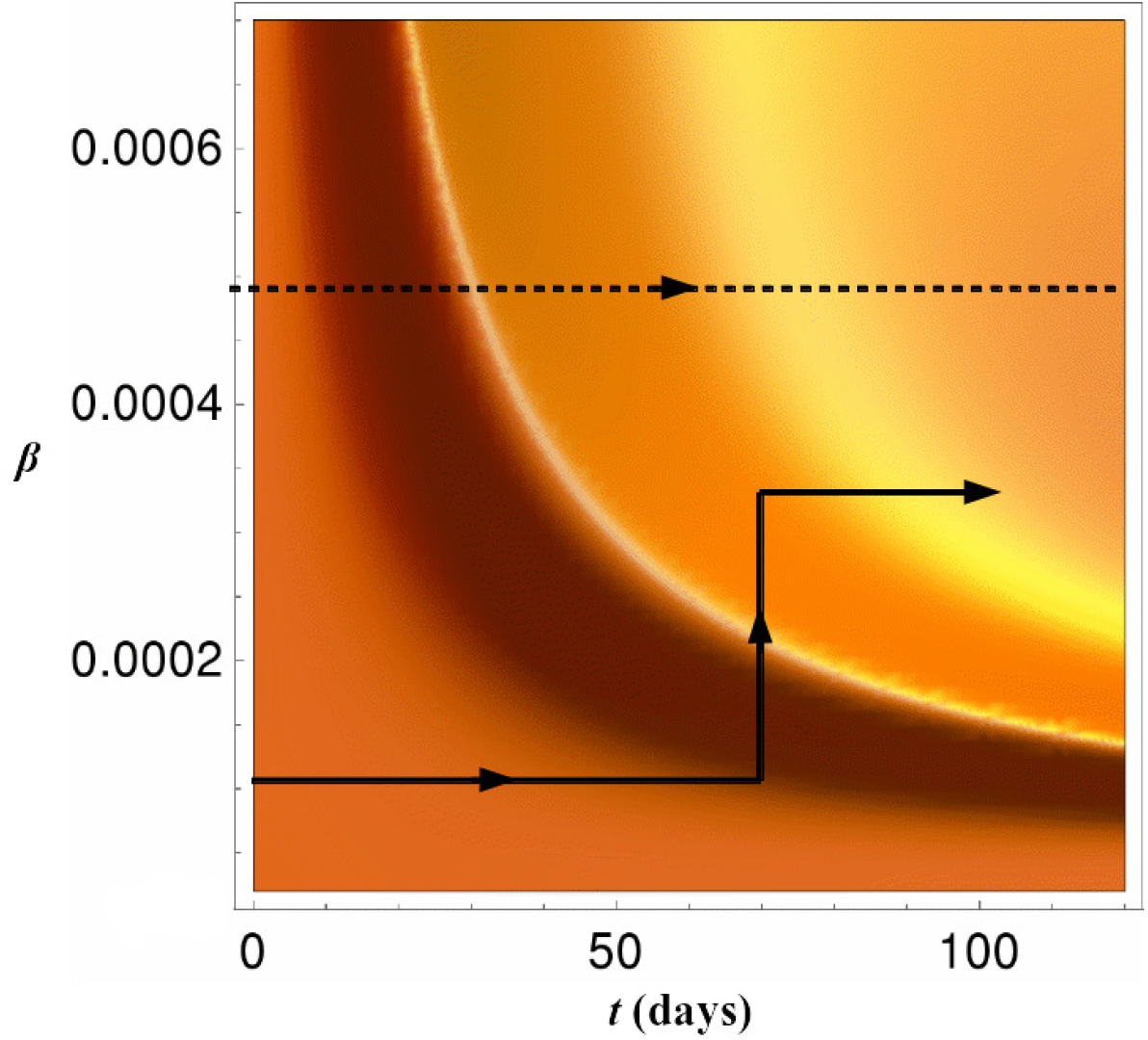
Top view of Fig. 5. Here the dashed line shows the estimated path of *I*(*t*) for the spring 2020 COVID-19 NYC epidemic through *I*(*β, t*) space. The lower path depicts a suggested route through which the intensity and time of the epidemic is shortened. *β* has units of days^−1^ per 1000 people.

Here the estimated path of *I*(*t*), using the value for *β* from the first row of Table 1 for the 2020 NYC epidemic, is shown by a dashed line. An interesting feature of this figure is that it can be seen that as the ridge bends for decreasing *β* the peak value for *I* is lowered but for a constant *β* path the epidemic is spread over a longer time period. That is, as *β* is lowered the constant *β* paths for *I*(*t*) begin to take a sideways and much longer route over the ridge.

This result suggests another possible approach to managing the epidemic. If *β* were to be decreased by aggressive mitigation then one might approach the ridge at a location where it is bent to the right in time. This level of mitigation is held in place until the ridge is near then if given a sudden increase in *β*, over a short time period, the epidemic will cross the ridge almost vertically in Fig. 7 at a point where it is greatly reduced in height. Once across, the epidemic is then completed at the newly increased constant value for *β*. A path of this sort is sketched in the lower section of Fig. 7. Therefore, within the framework of this model, a path of this nature greatly lessens the intensity and shortens the time period for the infected phase of the epidemic.

## Data Availability

All data used in this report is either listed in the report or can be found at the link below.

http://www1.nyc.gov/site/doh/covid/covid-19-data.page

